# Immersive 3D Visualisation (3D Printing, Virtual Reality) enhances spatial understanding of complex congenital heart defects

**DOI:** 10.1101/2024.05.05.24306500

**Authors:** Mahesh Kappanayil, Aishwarya Gurav, Sarin Xavier, Harikrishnan Anil Maya, Balaji Srimurugan, Abish Sudhakar, Raman Krishna Kumar

**Affiliations:** Department of Pediatric Cardiology, Amrita Institute of Medical Sciences, Kochi, Kerala, India; Cardiovascular 3D Printing and Extended Reality Laboratory, Amrita Institute of Medical Sciences, Kochi, Kerala, India; Department of Pediatric Cardiovascular and Thoracic Surgery, Amrita Institute of Medical Sciences, Kochi, Kerala, India

**Keywords:** : Cardiovascular imaging, Spatial Intelligence, congenital heart disease, 3D imaging, digital twin, extended reality

## Abstract

**Background:** Diagnosing, managing complex CHD demands excellent morphological understanding. Individual differences in visuospatial skills, training and experience can impact spatial interpretation of volumetric cardiac imaging. Immersive 3D visualisation may help overcome these challenges, but evidence of clinical benefit is lacking.

This study explores variability in visuospatial abilities and interpretation of conventionally viewed volumetric cardiac imaging data among members of a pediatric cardiac unit, and impact of using immersive 3D formats (3D-prints, Virtual Reality) on spatial understanding of complex CHD morphology.

**Methods:** Prospective cohort study involving 9 heterogenous members of an advanced pediatric cardiac program [3 consultant cardiologists, 2 cardiothoracic surgeons, 1 cardiac radiologist, 3 cardiology trainees]. Participants’ visuospatial abilities were quantified using a validated test (Revised PSVT:R). Understanding of spatial relationships between anatomical structures was assessed using structured questionnaires for 17 unique anonymised volumetric cardiac scans (15 CT, 2 MRI) of complex CHD visualised in three formats 1). conventional DICOM (CDICOM); 2). 3D prints (3DP); 3). Virtual Reality (VR). Accuracy, time taken, perceived level of difficulty, and confidence in interpretation were assessed and compared.

**Results:** Spatial abilities varied widely (median 8, IQR 6-30), independent of expertise/experience. Limitations in conventional reading were significantly overcome with immersive 3D. Mean accuracy score of 60.48% ±17.13% with CDICOM increased to 83.93% ± 7.84% with 3DP, and 90.81% ± 5.03% with VR (p<0.001). 3DP and VR permitted significantly faster interpretation (p<0.001), with significantly better ease and confidence. While immersive 3D visualisation led to significantly improved spatial understanding for all, it also minimised differences between participants with widely variable skill and experience levels.

**Conclusion:** Spatial abilities are variable. Immersive 3D visualisation can enhance spatial understanding of complex CHD morphology, overcoming challenges in spatial intelligence, experience, expertise. These technologies may be suitably leveraged as effective clinical and teaching tools in congenital cardiology.

**What is already known on this subject:** There is increasing exploration of use of novel immersive 3D technologies like 3D printing and Virtual/Augmented Reality in planning congenital cardiac surgery. Case reports and case series cite their use in visualising cardiovascular imaging data, but do not offer objective evidence or mechanistic insights on *how* immersive 3D interaction helps.

**What this study adds:** This study provides objective and subjective evidence that 3D printed and Virtual Reality representations of volumetric cardiovascular imaging data results in improved spatial anatomic understanding of complex cardiac defects among members of a pediatric cardiac care team. It also highlights variability in spatial intelligence and clinical experience among team members, and that immersive 3D can help overcome these challenges while interpreting cardiac imaging information.

**How this study might affect research, practice or policy:** Greater integration of immersive 3D visualisation tools in clinical practice may improve quality of care by improving physician-understanding of complex anatomical problems. It also makes a case for use of 3D printed and digital cardiac morphological specimens in training pediatric cardiac professionals.

## Introduction

Immersive three-dimensional (3D) technologies −3D Printing (3DP), Extended Reality (XR) - offer powerful new ways to engage with digital data by converting them into physical prototypes (3D prints) or computer-generated virtual 3D objects, environments (XR). Pediatric cardiac care may present transformative use-cases - spanning diagnosis, procedural planning, communication, teaching, research, simulation and collaboration.^1–7^ These technologies are still nascent; wider adoption awaits scientific validation.^4–7^

Pediatric cardiac professionals are challenged by need to diagnose and manage wide range of anatomical complexities. Most diagnostic cardiovascular imaging is either 2-dimensional (2D), *or* viewed in 2D formats. Volumetric datasets like CT, MR angiography are also typically viewed as 2D slices, or volume renderings on flat screens. Mentally reconstructing complex 3D anatomies from 2D images requires visuospatial abilities, training and experience. While skill and experience can be acquired, spatial intelligence, composed of various elements like abstract thinking, mental strategising, rotational understanding may be unique, variable trait.^8^ Variability in visuospatial abilities among pediatric cardiac professionals has not been studied. Immersive technologies may simplify visualisation and 3D understanding of complex morphologies, potentially compensating for variations in inherent and acquired skills.

This study explores variability in spatial intelligence, and impact of using immersive 3D technologies (3D printing, Virtual Reality) on spatial interpretation of tomographic cardiovascular images of complex CHD, among diverse members of a high-competency pediatric cardiac unit.

## MATERIALS AND METHODS

### i). Setting and Participants

Prospective observational study conducted over 20 months at a high volume, high expertise pediatric cardiac unit ^9–11^. Supported by dedicated cardiac radiology service - unit cardiologists, surgeons and trainees regularly view, interpret tomographic images in day-to-day clinical workflow. Complex management decisions are taken in joint multidisciplinary meetings. Unit has point-of-care Cardiovascular 3D Printing and XR Laboratory ^3^.

Nine team members volunteered to participate - six faculty (10-30 years of individual domain experience) including three pediatric cardiology consultants (PCC), two pediatric cardiac surgeons (PCS) and one cardiac radiologist (CR) - and three pediatric cardiology fellows (PCF) (1 month to 2 years into training).

### ii). Test of Spatial Abilities

All participants underwent test of spatial abilities - Revised Purdue Spatial Visualization test - Test of Rotations [Revised PSVT:R] (with permission from author) ^12^. Administered in standardised manner, test required subjects to answer thirty questions within twenty minutes, assessing ability to interpret change in visual perspective upon rotation of 2-dimensional diagrams of 3-dimensional shapes. Responses were tallied against correct answers provided in answer-key - assigned score of 1 for correct and nil for incorrect responses - scores were totalled (n/30).

### iii). Selection and Preparation of Test-case Tomographic Imaging Datasets

17 tomographic datasets (CT, MRA) of complex CHD cases were selected from institutional database (preceding ten years) after screening for data quality (resolution, slice thickness, readability, suitability for conversion to 3D formats). Cases included mix of spatially complex cardiovascular anomalies; situs and cardiac position variations were also represented. (Table 1).

**Table 1:**
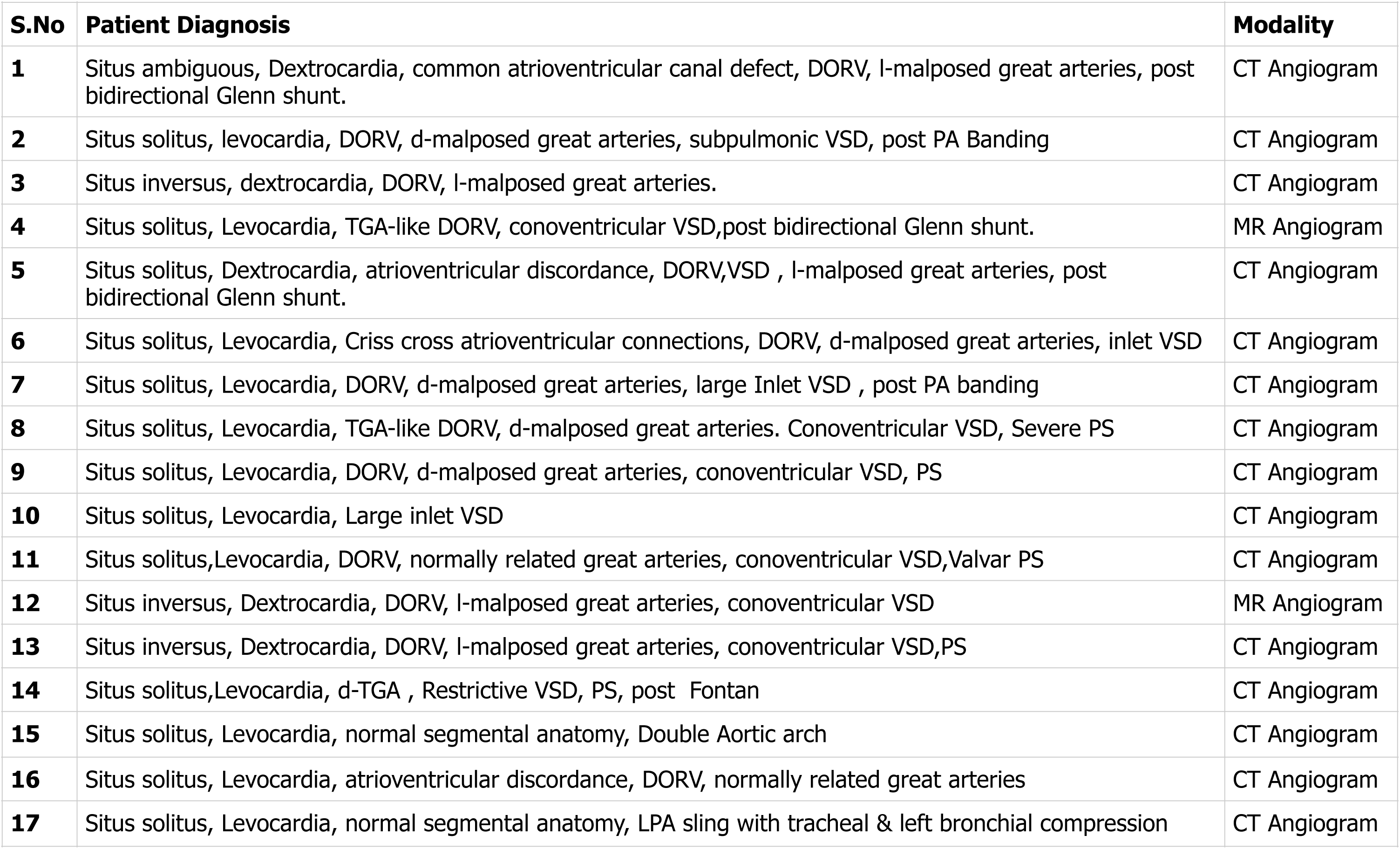
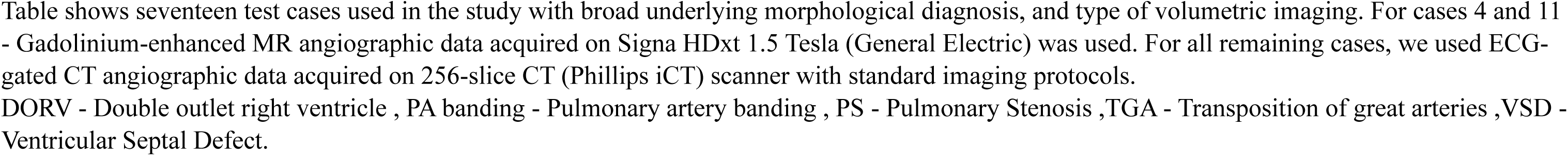
Test-cases - basic morphological diagnosis and imaging modality.

Clinical data, echocardiography, operation notes (where available) and other relevant information were thoroughly reviewed. Case-specific questionnaires were prepared with 5 to 7 multiple-choice questions each assessing ability to identify spatial relationships between anatomical segments - e.g., atrioventricular relationships, spatial positioning of cardiac chambers and great arteries, locations of ventricular septal defect etc. Case-specific answer keys were prepared.

### iv). Visualisation Formats

Test datasets were anonymised, prepared for three visualisation formats : i.conventional DICOM (CDICOM); ii.3D printed (3DP) models; iii.Virtual Reality (VR).

*Conventional* visualisation was using standard workstation-based DICOM viewing platform (Horos, Horos Project, USA) which all participants were already conversant with, and included in-built tools like multi-planar viewing, windowing, thresholding, sizing, measuring etc. No special training was required.

#### Conversion to Immersive 3D Formats

3D conversions were done by expert team at in-house 3D Labs. DICOM data was imported into proprietary software (Materialise Innovation Suite V26.0, Materialise NV, Belgium). Standard workflow was followed for segmenting region-of-interest and converting to digital 3D file format (.STL) ^4,13^. ‘Hollow’ models with appropriate cut-planes were designed for intracardiac visualisation, and ‘solid’ models for predominantly extracardiac anomalies. Markers of orthogonal planes were incorporated. STL files were digitally optimised (Preform, Formlabs, USA) and 3D printed in 1:1 scale on Form2 3D printer (Formlabs, USA), using clear photopolymer resin. Models were post-processed, and optimised for administering.

*VR visualisations* were done using software applications - FDA-approved 3D medical modelling software Elucis (Realize Medical, Canada), 3D volume-rendering software Vea (Vea Simulations, Poland) and homegrown VR visualisation platform (Unity Game Engine, Unity Technologies). VR tools included interactions like grabbing, scaling, cutting, labelling, dissecting, annotating (using handheld ‘game-controllers’). Hardware included Meta Quest2 (Meta, USA) and HTC Vive-Pro (HTC, Taiwan) VR headsets tethered to GPU workstation. Participants were trained for VR using dummy datasets, until adequately proficient.

### v). Tests

Test-cases were administered to all participants in all three formats in random, shuffled manner. Participants answered case-specific questionnaires (identical across visualisation formats), blinded to clinical and other imaging information. Sessions were timed with stopwatch.

Participants were scored for accuracy (correct=1, incorrect=0) and time taken (seconds). 10-point Likert’s scale was used to assess subjective elements - i). perceived level of difficulty (1 - ‘very easy’ and 10 - ‘very difficult’). ii). level of confidence in interpretation (1 - ‘not confident’, 10 - ‘very confident’) ^13^.

Mean values were calculated for individual participants for each visualisation mode across all test datasets. Subgroup comparisons were made between cardiologists, surgeons, fellows and cardiac radiologist. Comparisons were also made between experienced pediatric cardiac specialists (PCC plus PCS) and fellows-in-training.

### v). Statistical Analysis

Pearson Chi-Square test was used to compare categorical variables. Spearman’s correlation was used to find the correlation between spatial orientation score and accuracy scores and time across the three visualisation modalities. Paired t-test was used to compare the continuous variables among participants across modalities. A repeated measures ANOVA with Post hoc Bonferroni cor-rection was used to compare accuracy score and time in different modalities for participants. Statistical analyses were conducted using SPSS Version 20.0 for Windows (IBM Corporation ARMONK, NY, USA).

## RESULTS

*All 9 participants completed spatial ability test and analysis of 17 imaging datasets across test visualisation formats. 459 test-case questionnaires were analysed. Tables 2 and 3 summarise accuracy scores and time-performance*.

**Table 2:**
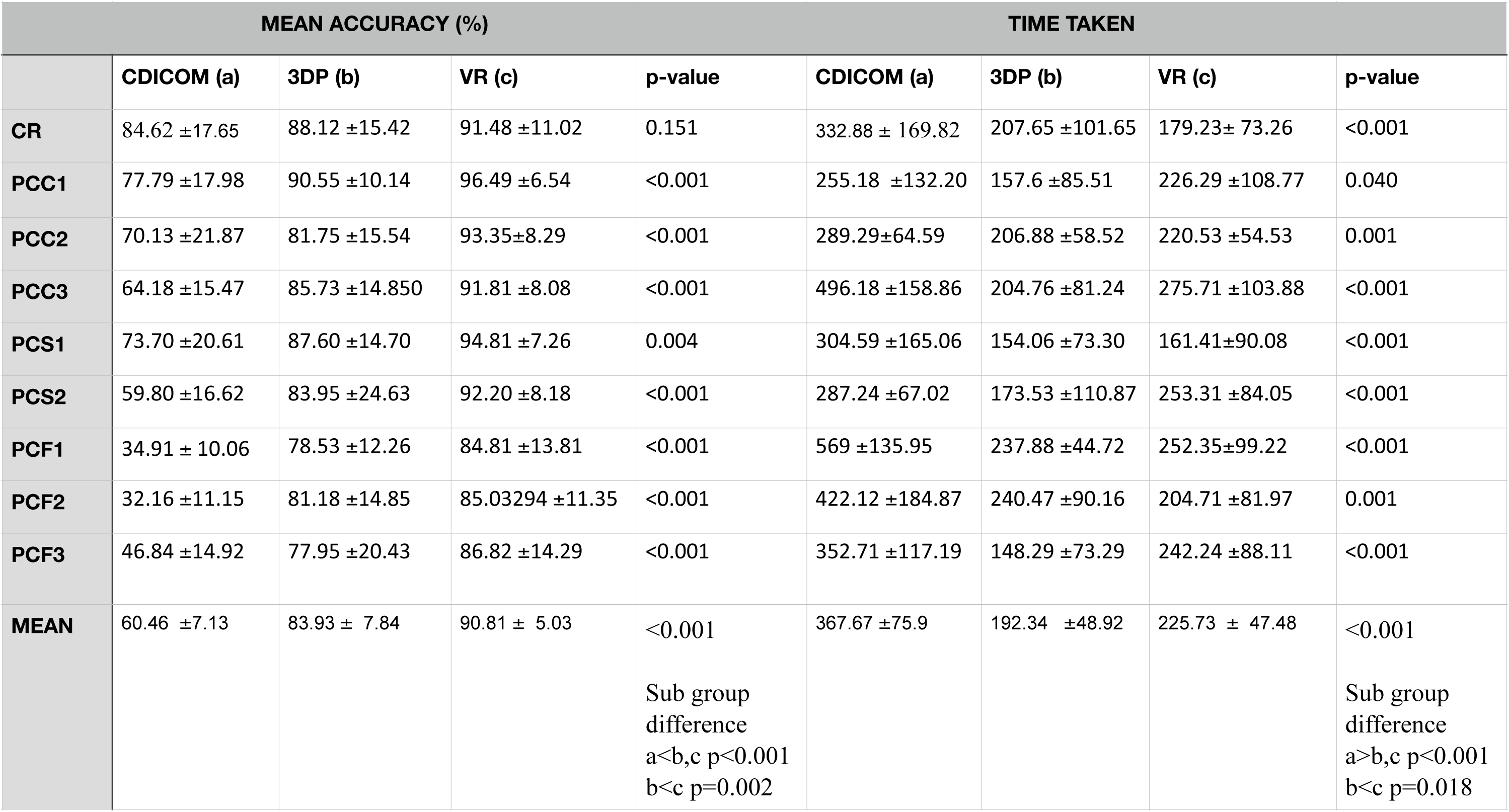
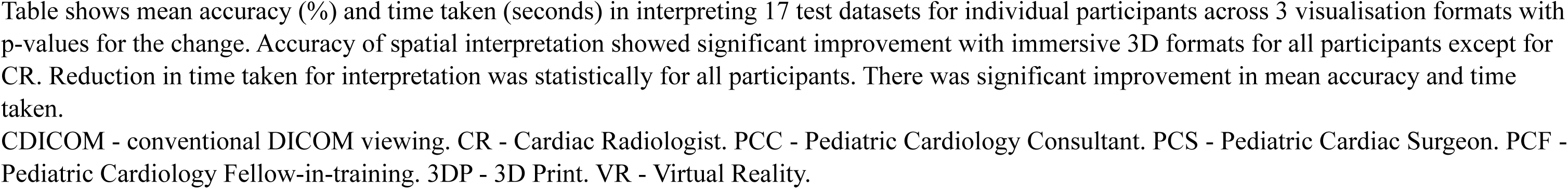
Mean Accuracy (%) scores and time taken (seconds) - individual participants.

**Table 3.** Mean Accuracy (%) scores and time taken (seconds) - participant subgroups.

|  | Category | CDICOM (a) | 3DP (b) | VR (c) | P Value | Subgroup Difference |
| --- | --- | --- | --- | --- | --- | --- |
| ACCURACY SCORE (%) | CR | 84.62 ± 17.65 | 88.11 ± 15.4 | 91.48 ± 11.02 | 0.151 | NA |
|  | PCC | 70.7 ± 11.2 | 86.02 ± 9.14 | 93.9 ± 5.5 | <0.001 | a<b, c p<0.001<br>b<c p=0.010 |
|  | PCS | 66.8 ± 13.21 | 85.77 ± 14.1 | 93.7% ± 6.6 | <0.001 | a<b, c p<0.001<br>b<c p=0.078 |
|  | PCF | 37.95 ± 7.08 | 79.2 ± 8.96 | 85.8 ± 6.7 | <0.001 | a<b, c p<0.001<br>b<c p=0.017 |
|  | p-value | <0.001 | 0.06 | 0.006 |  |  |
| TIME TAKEN (SECONDS) |  |  |  |  |  |  |
|  | CR | 332.88 ± 169.82 | 207.647 ± 101.65 | 179.23 ± 73.26 | <0.001 | a>b p=0.006<br>a>c p=0.011<br>b>c p=0.973 |
|  | PCC | 346.86 ± 74.89 | 189.72 ± 59.4 | 240.84 ± 60.0 | <0.001 | a>b, c p<0.001<br>b<c p=0.034 |
|  | PCS | 295.92 ± 96.8 | 163.79 ± 76.01 | 207.50 ± 69.15 | <0.001 | a>b, c p<0.001<br>b<c p=0.033 |
|  | PCF | 447.94 ± 112.25 | 208.8 ± 36.98 | 238.27 ± 58.18 | <0.001 | a>b,c p<0.001<br>b<c p=0.089 |
|  | p-value | 0.002 | 0.080 | 0.028 |  |  |

Table shows mean accuracy (%) and time taken (seconds) in interpreting test datasets for participant subgroups across 3 visualisation formats, with respective p-values for the change. There was statistically significant difference in accuracy scores between the four subgroups while interpreting conventional (CDICOM) and VR formats, but not with 3D prints. All subgroups except CR showed significant improvement with immersive 3D formats. Improvement in speed of interpretation (time taken) was significant for all subgroups with 3D formats. While difference between subgroups was highly significant with conventional, it was reduced with use of VR and insignificant with 3D prints.
CDICOM - conventional DICOM viewing. CR - Cardiac Radiologist. PCC - Pediatric Cardiology Consultants. PCS - Pediatric Cardiac Surgeons. PCF - Pediatric Cardiology Fellows-in-training. 3DP - 3D Print. VR - Virtual Reality.

### Spatial Abilities

Revised PSVT:R scores showed wide variability ranging from 0-15/30, median score 8 (IQR 6.5-30) (figure 1), independent of experience or expertise.

**FIGURE 1:**
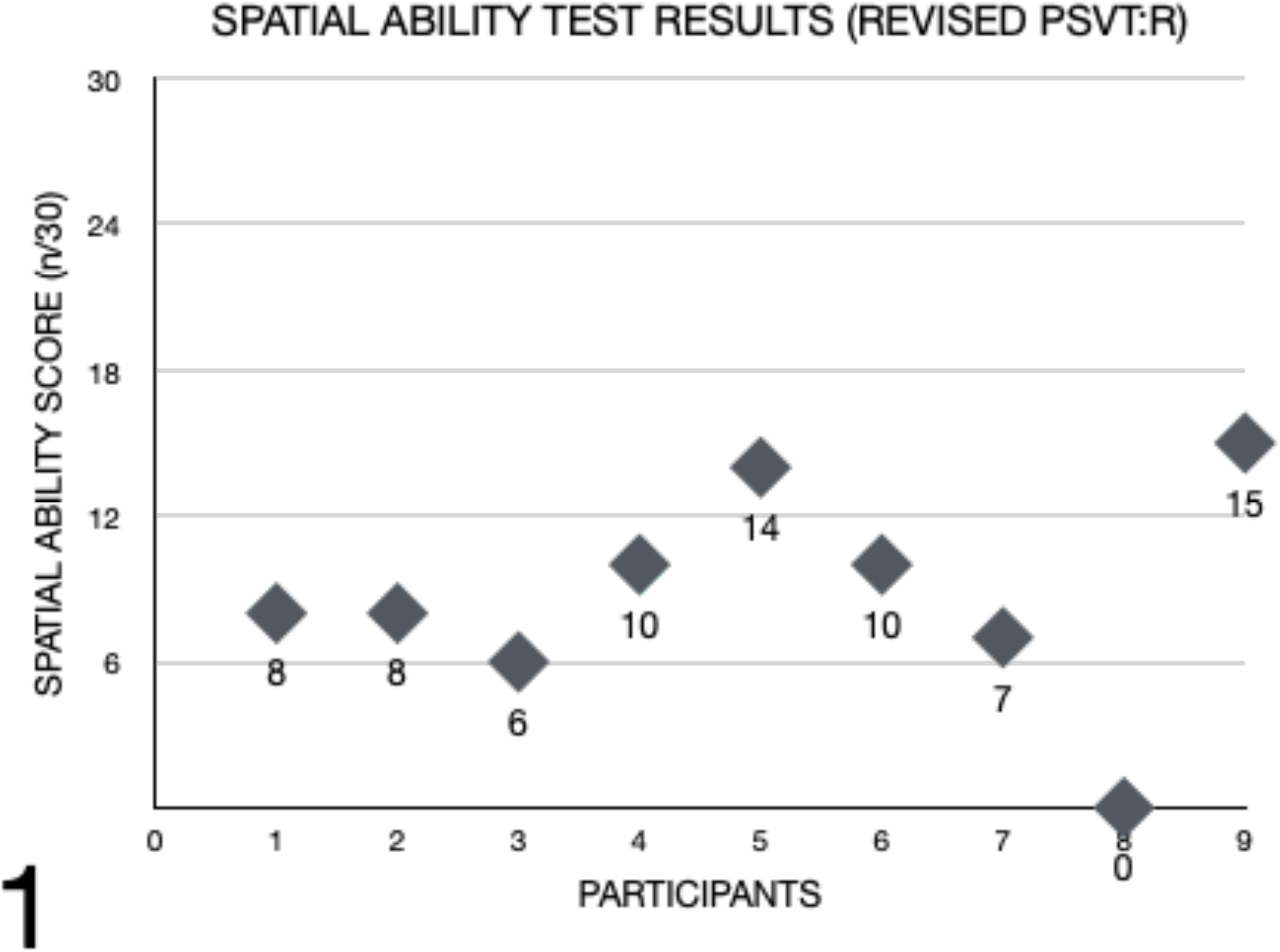
Spatial Intelligence. Scatter diagram showing spatial ability scores of individual participants on Revised Perdue Spatial Visualisation Test - Test of Rotations (PSVT:R).

### Accuracy

Conventional reading had lowest accuracy, which improved with 3DP and VR (Table 2) - statistically significant for all except cardiac radiologist. Mean accuracy score with CDICOM (60.46%±7.13%) improved to 83.93% ± 7.84% with 3DP, and to 90.81% ± 5.03% with VR (p<0.001). Improvement from 3DP to VR was statistically significant (p=0.002). (Table 2).

Table 3 compares accuracy scores of subgroups across formats, as well as difference in accuracy within each format across participant groups.

#### i). Accuracy across formats

Change in accuracy scores is shown in figure 2A - improved from CDICOM through 3DP to VR, with change being statistically significant for consultant cardiologists (p<0.001), surgeons (p<0.001) and fellows (p<0.001) (table 3).

**FIGURE 2:**
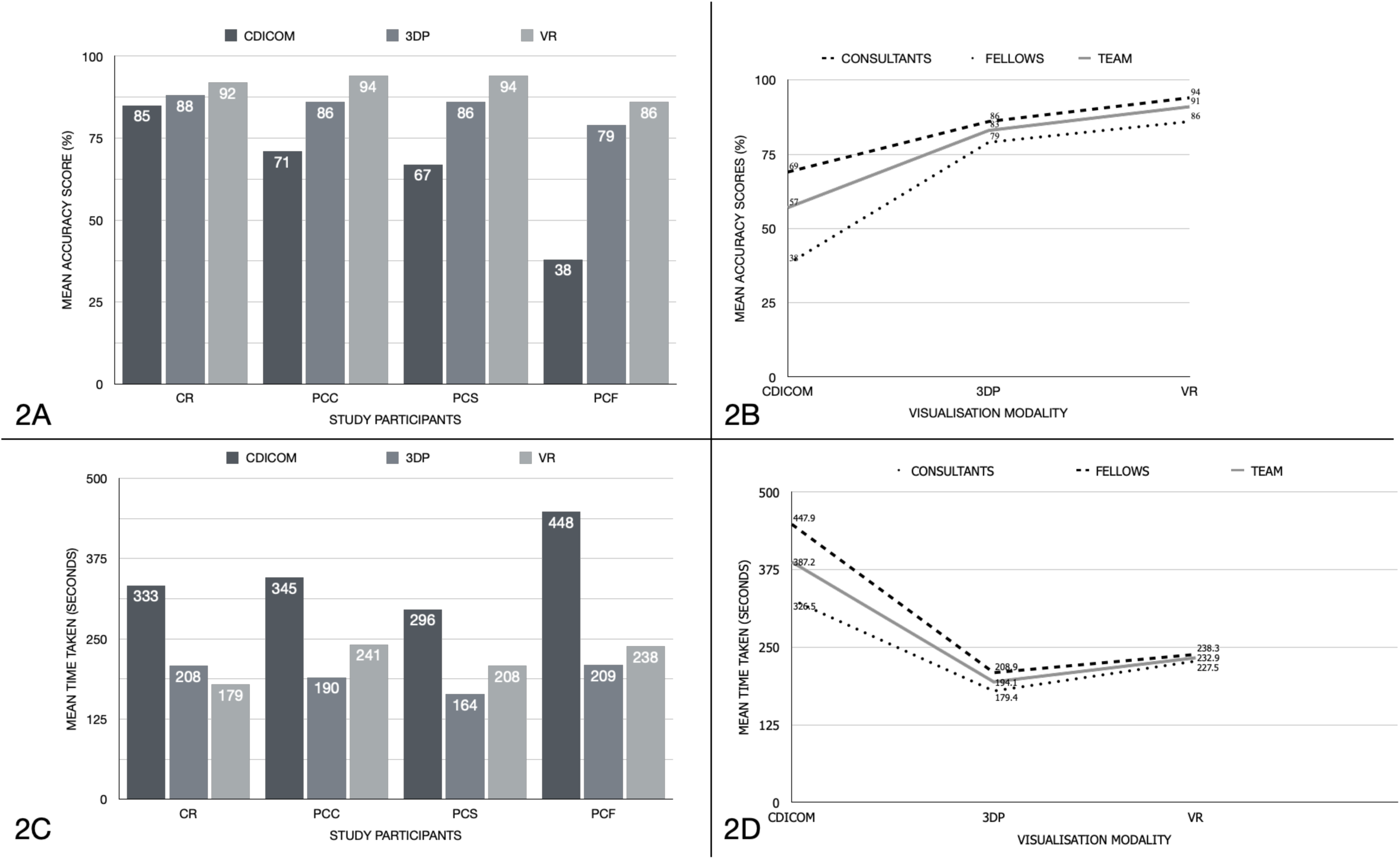
Accuracy and TimeTaken. **2A** : Mean accuracy scores (%) of participant subgroups in interpreting 17 test-cases using three visualisation formats. Accuracy improved for all subgroups with immersive visualisation - VR en-abled best accuracy. **2B** : Comparison of mean accuracy scores of pediatric cardiac fellows (dotted line) with consultants (dashed line) - wide gap with conventional visualisation significantly narrowed with 3D print and VR. Solid line represents mean scores for composite pediatric cardiac team. **2C** : Mean time taken (seconds) for interpretation - significant reduction with immersive 3D for-mats. 3D prints required least time except for cardiac radiologist (VR fastest). **2D** : Comparison of time taken by fellows (dotted line) and consultants (dashed line) - wide gap with conventional, narrowed with 3D print and VR. Solid line represents mean combined scores. CDICOM - conventional DICOM. CR - cardiac radiologist. 3DP −3D print. PCC - pediatric cardi-ology consultants. PCS - pediatric cardiac surgeons. PCF - pediatric cardiology fellows. VR - virtu-al reality.

#### ii). Accuracy across participant groups

##### CDICOM

Cardiac radiologist performed best at conventional visualisation with mean accuracy score of 84.62% ± 17.65%, followed by cardiology consultants (70.7% ± 11.2%), surgeons (66.8% ± 13.21%) and fellows (37.95% ± 7.08%). Difference in accuracy scores was statistically significant (p<0.001). Mean accuracy score for ‘pediatric cardiac team’ (PCC + PCS + PCF) was 57.43% ± 7.15 %. Gap between pediatric cardiology fellows (PCF) and consultants (PCC + PCS) was wide (37.95% ± 7.08% vs 69.12± 9.14 %, p<0.001).

##### 3DP

Mean accuracy scores significantly improved for all with 3D prints (Tables 2,3; Figure 2A), resulting in statistically insignificant difference between them (p=0.06) (Table 3). Pediatric cardiac team mean score improved to 83.41% ± 7.29 %. Difference between fellows and the consultants narrowed (79.2% ± 8.96 % vs 85.92% ± 9.12 %), though a statistically significant difference between the two persisted (p=0.02).

##### VR

Best accuracy was with VR (Tables 2,3; Figure 2A) - mean score of pediatric cardiac team 90.72% ± 5.04 %. Though there was statistically significant difference between accuracy scores of fellows (85.55± 6.71 %) and consultants (93.82 ± 5.25 %) with VR (p<0.001), there was marked improvement from conventional, with impressive narrowing of gap.

Figure 2B summarises change among pediatric cardiac consultants and fellows. Comparative accuracy of pediatric cardiac consultants and fellows while analysing specific elements of spatial anatomy is shown in supplemental file.

###### 2. Time Taken

CDICOM interpretation took significantly longer time than 3DP and VR (Table 2).

Figure 2C compares time taken by participant subgroups - significantly lesser time required with 3DP and VR (p<0.001) (Table 3). Consultant cardiologists and surgeons were faster with 3DP than with VR (p=0.034 and p=0.033 respectively). Fellows were also faster with 3DP than VR, but difference was not statistically significant (p=0.089). Radiologist took longer to interpret 3DP than VR; difference not statistically significant (p=0.973).

Difference in time taken by four subgroups was statistically significant while interpreting CDICOM (p=0.002) and with VR (p=0.028), but not for 3DP (p=0.080) (Table 3). There was wide, statistically significant difference between cardiac consultants (326.48± 69.94 seconds) and fellows (447 ± 94 seconds), p<0.001 with CDICOM; gap narrowed with immersive 3D (Figure 2D). There was no significant difference between them while interpreting VR (227.51± 53.37 vs 238.27± 58.18 seconds, p=0.375); borderline significance remained with 3DP (179.35± 60.94 vs 208.88± 36.98 seconds, p=0.047).

###### 3. Subjective Parameters

Table 4 compares subjective perceptions of ‘level of difficulty’ and ‘level of confidence’.

**Table 4 :** Subjective scores of ‘level of difficulty’ and ‘level of confidence’ while interpreting the three test modalities across participant groups.

|  |  | CDICOM (a) | 3DP (b) | VR (c) | P Value | Difference |
| --- | --- | --- | --- | --- | --- | --- |
| PERCEIVED LEVEL OF DIFFICULTY<br>(LIKERT’S SCALE OF 0-10) | CR | 5.94 ± 2.36 | 4.94 ± 2.01 | 3.00 ± 1.27 | <0.001 | a>b p=0.209<br>a>c p=0.001<br>b>c p=0.002 |
|  | PCC | 5.03 ± 1.00 | 3.94 ± 1.088 | 3.2 ± 0.97 | <0.001 | a>b p=0.005<br>a>c p<0.001<br>b>c p=0.095 |
|  | PCS | 4.21 ± 0.81 | 3.47 ± 0.67 | 3.47 ± 1.15 | 0.004 | a>b p=0.008<br>a>c p<0.027<br>b=c p=1.000 |
|  | PCF | 7.7 ± 0.904 | 4.47 ± 0.858 | 3.1 ± 0.567 | <0.001 | a>b,c p<0.001<br>b>c p<0.001 |
|  | p-value | <0.001 | 0.011 | 0.319 |  |  |
| PERCEIVED LEVEL OF CONFIDENCE<br>(LIKERT’S SCALE OF 0-10) | CR | 8.12 ± 1.00 | 7.76 ± 1.64 | 8.17 ± 0.88 | 0.405 | NA |
|  | PCC | 7.13 ± 0.67 | 7.80 ± 0.745 | 7.84 ± 0.81 | 0.001 | a<b p=0.005<br>a<c p=0.006<br>b<c p=1.000 |
|  | PCS | 6.73 ± 0.73 | 7.91 ± 0.51 | 7.29 ± 1.03 | <0.001 | a<b p<0.001<br>a<c p=0.051<br>b>c p=0.048 |
|  | PCF | 3.66 ± 0.62 | 6.67 ± 0.858 | 7.43 ± 0.60 | <0.001 | a<b p<0.001<br>a<c p<0.001<br>b<c p=0.006 |
|  | p-value | <0.001 | 0.007 | <0.001 |  |  |

Table shows mean ratings on 10-point Likert scale for perceived level of difficulty and level of confidence of participant subgroups while interpreting the test imaging datasets across the three visualisation formats, with respective p-values for significance.

###### Level of difficulty

Conventional was rated most difficult, VR simplest to interpret (p<0.001). There was statistically significant difference between subgroups for CDICOM (p<0.001) and for 3DP (p=0.011), but not for VR (p=0.319). Reduction in difficulty ratings from CDICOM to 3DP to VR was statistically significant for all subgroups. Changes are shown in figure 3A.

**FIGURE 3:**
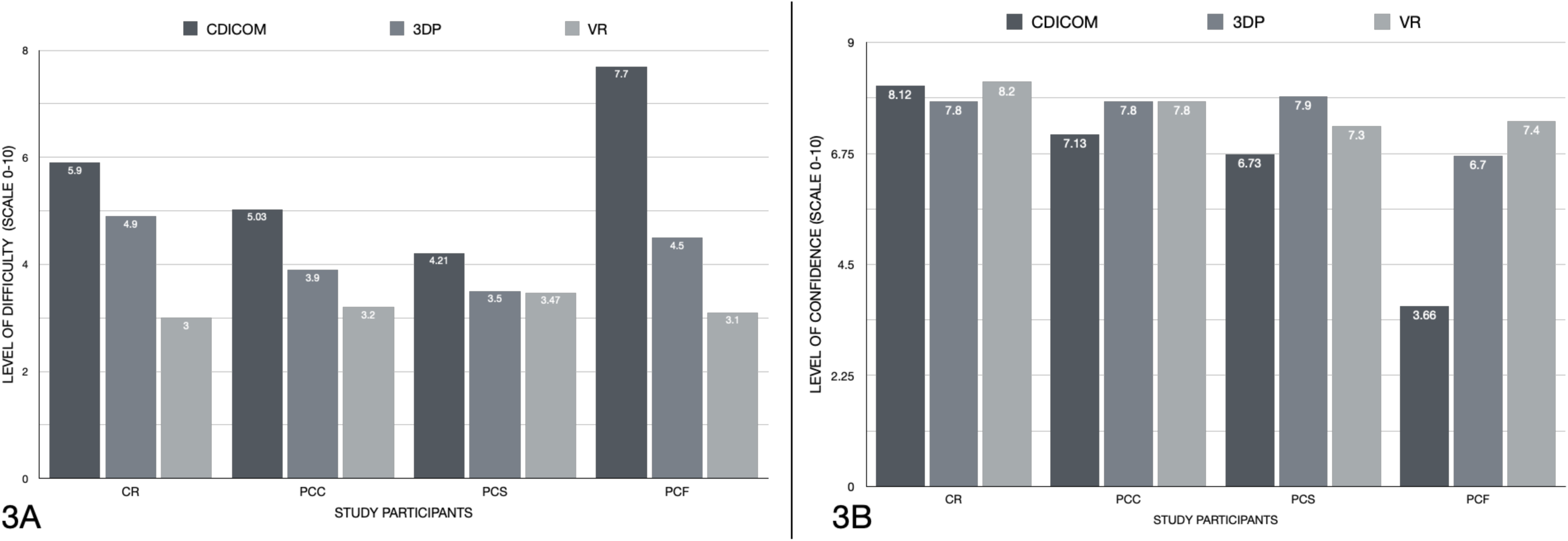
Subjective parameters [10-point Likert Scale]. **3A** : ‘Level of difficulty’ in interpretation perceived by participant subgroups using test formats - significant reduction with immersive 3D formats, VR perceived simplest. Change was most notable for fellows. **3B** : ‘Level of confidence’ in interpretation - improved with 3D prints and VR - most notable for the fellows. CDICOM - conventional DICOM. CR - cardiac radiologist. 3DP −3D print. PCC - pediatric cardi-ology consultants. PCS - pediatric cardiac surgeons. PCF - pediatric cardiology fellows. VR - virtu-al reality.

###### Level of confidence

While there was no significant change for cardiac radiologist, there was significant improvement for other three groups with immersive 3D. Surgeons had higher confidence with 3DP over VR (p=0.048), cardiology fellows were more confident with VR than 3DP (p<0.006). There was no statistically significant difference between VR and 3DP for consultant cardiologists. Changes are depicted in figure 3B.

Figure 4 and supplemental material (video) show examples of multi-format visualisations in the study.

**FIGURE 4:**
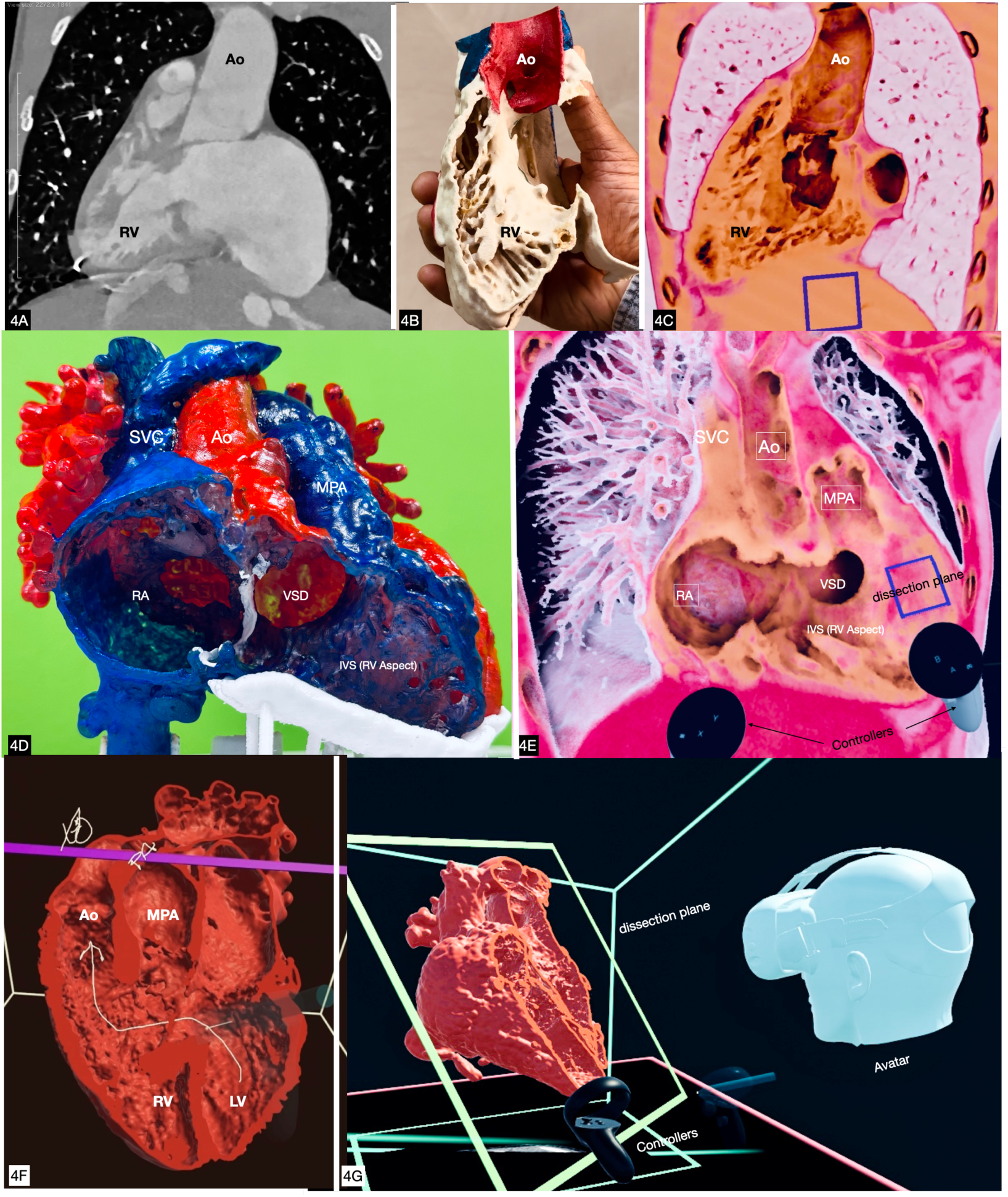
Visualisation modes. **4 A,B,C :** Computed tomography scan data of case of situs ambiguous, dextrocardia, common atri-oventricular canal defect, double outlet right ventricle, pulmonic stenosis as visualised in conven-tional format (4A), 3D printed model (4B) and VR (4C). **4D, E** : case of double outlet right ventricle, dextro-malposed Aorta, large inlet VSD with conoven-tricular extension. **4D** −3D printed model with anterior walls subtracted to show intracardiac anatomy. **4E** - intracardiac anatomy with VR virtual dissection, coronal plane. **4F** : intracardiac anatomy of case of double-outlet right ventricle in VR, virtual annotation of LV-to-Aorta pathway (white arrow). **4G** : VR session using digital ‘dissection plane’, operator’s position represented by ‘avatar’. *VR software* : 4C,E [Vea Simulations, Poland]. 4F,G [Elucis, Realize Medical, Canada] Ao - aorta. Controllers - handheld controls. MPA main pulmonary artery. IVS - interventricular septum. LV - left ventricle. RA - right atrium. RV - right ventricle.

##### Spatial abilities & accuracy, speed of interpretation

Spatial ability score had weak positive correlation with accuracy, weak negative correlation with speed of interpretation of CDICOM - statistically insignificant. Similarly, statistically insignificant weak positive correlations were noted between spatial ability score and accuracy at interpreting 3DP and VR. Spatial ability score had a strong negative correlation with time taken for interpreting 3DP (p=0.002); correlation was weak, statistically insignificant for VR (p=0.317).

## DISCUSSION

Immersive 3D technologies can transform visualisation and interaction with medical imaging data. This study, for first time, explores differences in visuospatial abilities among members of a pediatric cardiac team, and impact of immersive 3D visualisation on their interpretation of volumetric imaging.

Pediatric cardiac professionals have been among earliest healthcare adopters. 3D printed heart models have been increasingly used in procedural planning in recent years ^1–4,7,14–16^. More recently, XR is being explored for digital immersion capabilities ^5–7^. Prime driver has been need for better morphological understanding of complex CHD.

Conventional imaging and training do not adequately foster spatial understanding. Even ‘3D vol-ume-rendered’ images viewed on 2D workstation screens do not adequately convey depth and scale. Access to morphological specimens is rare - learning occurs mostly though textual descriptions, diagrams, photographs and accumulated experience. Nomenclatures and classifications aid identification, categorisation and communication ^17^. However, no two patients are exactly alike; appreciat-ing anatomical uniqueness is critical to management. Cardiologists are often straitjacketed into interpreting predefined ‘views’ and slices of imaging data, without conscious effort, opportunity and sometimes innate ability to adequately understand spatial anatomy. Complex variants of double outlet right ventricle, anatomically corrected malposition of great arteries, criss-cross hearts, ‘topsy-turvy hearts’, heterotaxies are examples of morphologies that can be challenging to spatially understand. Despite long learning curves, it is not uncommon to be repeatedly surprised by morphological complexities not encountered previously. Errors in understanding can impact management decisions and outcomes.

‘Spatial intelligence’ is mental computational capacity to solve spatial problems, and to mentally retrieve, process, retain and transform visual information ^8,12^. Importance has been recognised in numerous fields including art, and STEM (Science Technology Engineering Mathematics) disciplines ^18^. Multi-dimensional in construct, spatial intelligence has at least two core aspects - spatial visualisation (SV) and spatial relation/orientation (SR).*’Mental rotational ability’* is the component that enables visualisation of a 3D object from different perspectives/viewpoints by mentally rotating it. Specialised tests, instruments have been developed for objective assessment ^12,19–21^. Purdue Spatial Visualisation Test (PSVT), with three subtests (‘Developments’, ‘Rotations’, ‘Views’), was originally developed by Guay in 1976; ‘Rotations’ subtest was subsequently extended to PSVT:R (Visualisation of Rotations) to test mental rotational ability, further modified by Yoon (2011) into Revised PSVT:R used for this study ^21^.

Limited studies have explored importance of visuospatial abilities among physicians ^22–25^. 3D understanding has been shown to be associated with anatomy learning among medical students^22–24^.Variability has also been shown to impact surgical skill acquisition and performance - especially in early stages of learning ^25^. While accumulation of experience and practice may obviate impact to some extent, lower spatial abilities may translate into longer learning curves, and need for better training techniques. Differences in spatial abilities among pediatric cardiac professionals has not been studied previously.

Immersive 3D technologies have potential to change the paradigm by presenting volumetric imaging data in spatial, interactive formats. 3D printing involves creation of physical prototypes by adding material layer-by-layer based on a digital design derived from patient-specific volumetric scans using computer-aided design (CAD) software ^2–4,7,15^. These 1:1 scale physical replicas provide uniquely immersive and tactile interaction, in sharp contrast to conventional viewing ^1–4, 7, 14–16^. Printed in suitable materials, they can be used for procedural simulation, communication, teaching,and even educating patients and families about upcoming procedures ^4,7,15^. 3D printing needs dedi-cated physical space, equipment, materials, software, and expertise - adoption has been restricted to select centres ^7^.

Extended Reality is an umbrella term encompassing VR, augmented reality (AR), and mixed reality (MR). While VR immerses users in a completely computer-generated environment, AR overlays digital information onto real world; MR merges elements of both. XR is typically experienced using specialised headsets or eyewear, software and handheld (or gesture) controls. Growth in digital technology especially 3D gaming tech has made XR increasingly accessible and affordable. XR offers powerful new means for immersive visualisation of volumetric cardiovascular imaging data ^5–7^. Incorporation of digital tools like virtual dissection, measurement, annotation, scaling, remote collaboration, and procedural simulation can further enhance interaction; 3D printed models have limited versatility and range of interactions. XR needs less infrastructure and turnaround times than 3D printing. On the downside, XR may be associated with cybersickness, vertigo and headache in some users ^5^.

Rising interest in their transformative potential is tempered by hesitancy in adopting these novel technologies in clinical workflows. Published literature, comprising predominantly case reports and case series, lacks clear evidence/validation of outcome benefits in CHD management.^4–7, 15,16^. This study was therefore designed to address a fundamental question - whether immersive 3D technologies can improve physicians’ understanding of complex CHD - as precursor to better clinical care and outcomes.We also tried to understand differences in spatial understanding between variably skilled/experienced individuals in a clinical setting, and potential to overcome these using immersive 3D visualisations. Supplemental figure graphically summarises the study.

### Test of Spatial Ability

Variability in Revised PSVT:R scores highlights heterogeneity of spatial abilities among physicians comprising a typical pediatric cardiac team - pointer towards need to develop better training, visualisation tools. Assessing true impact of variability, and potential determinants (gender, race, environment etc) would require larger, systematic studies.

### Accuracy of interpretation

Salient observations included significant limitations and variability in accuracy of interpretation with conventional visualisation (tables 2,3). Those with specialised training (cardiac radiologist - mean score) and/or with more experience (pediatric cardiac consultants) fared significantly better than trainees. Expectedly, cardiac radiologist with combination of training and experience fared best. Fellows and consultants were widely separated.

3DP and VR led to significant change with mean scores improving to 83.93% ± 7.84% with 3DP, and 90.81% ± 5.03% with VR, and remarkable reduction of gap between subgroups . Most impressive change was among trainees, bringing them near par with vastly more experienced senior consultants. With 3D printed models, there was no statistically significant difference between accuracy scores of consultants and trainees. With VR, even though there was statistically significant difference (p=0.006), the gap was markedly narrowed (table 3, figure 3).

### Time Taken

Immersive 3D visualisation permitted faster interpretation. Significant differences between sub-groups (especially consultants and fellows) with conventional format, were markedly reduced with 3DP and VR (p<0.001). Time difference between groups was insignificant with 3DP (p=0.080), with quickest interpretation for all except cardiac radiologist (VR fastest). Interestingly, while VR allowed best accuracy, it required more time than 3DP for most - possibly due to immersive, ‘gami-fied’ engagement in VR as also need to adapt to unfamiliar hardware (headsets, controllers).

### Subjective parameters

Immersive 3D formats simplified spatial understanding for all participants, reflecting in significantly lower scores for ‘level of difficulty’ and higher scores for ‘level of confidence’. Gains were again most notable for trainees, reducing gap between them and the more experienced consultants.

## Conclusions

**S**tudy offers novel insights into variability in spatial abilities, and interpretation of volumetric imaging of complex CHD among members of a real-world pediatric cardiac team. It provides objective evidence that immersive 3D visualisation can help overcome multi-dimensional individual variabilities, improving understanding and minimising gaps - equipping physicians for better, safer and more precise patient care. Immersive learning tools and collaborations based on these novel technologies may shorten learning curves and elevate quality of pediatric cardiac care.

## Limitations

Single center study with limited number of participants, test cases. Imaging data was archival, and clinical decisions, outcomes were not analysed. 3D formats were created at in-house 3D Printing, XR Lab - capabilities unavailable at majority of centres.

## Data Availability

All data produced in the present study are available upon reasonable request to the authors

## Abbreviations

CHD: Congenital heart disease
CT: computed tomography
DICOM: Digital Imaging and Communication in Medicine
MRI: magnetic resonance imaging
PSVT:R: Perdue Spatial Visualisation Test - Test of Rotations

## Ethics

Study was approved by Institutional Ethics Committee.

## Source of funding

Study was funded by institutional research grant. No funding from external agencies or industry.

## Disclosures

None

## SUPPLEMENTAL FILE (FIGURE) : Central Illustration

Interpreting cardiovascular cross-sectional imaging using conventional viewing formats requires training, experience and spatial abilities. Use of immersive 3D visualisation formats (3D prints, Extended Reality) can help overcome challenges, allowing more accurate, faster and easier interpretation, shortening learning curves and reducing differences among diverse members of cardiac team.

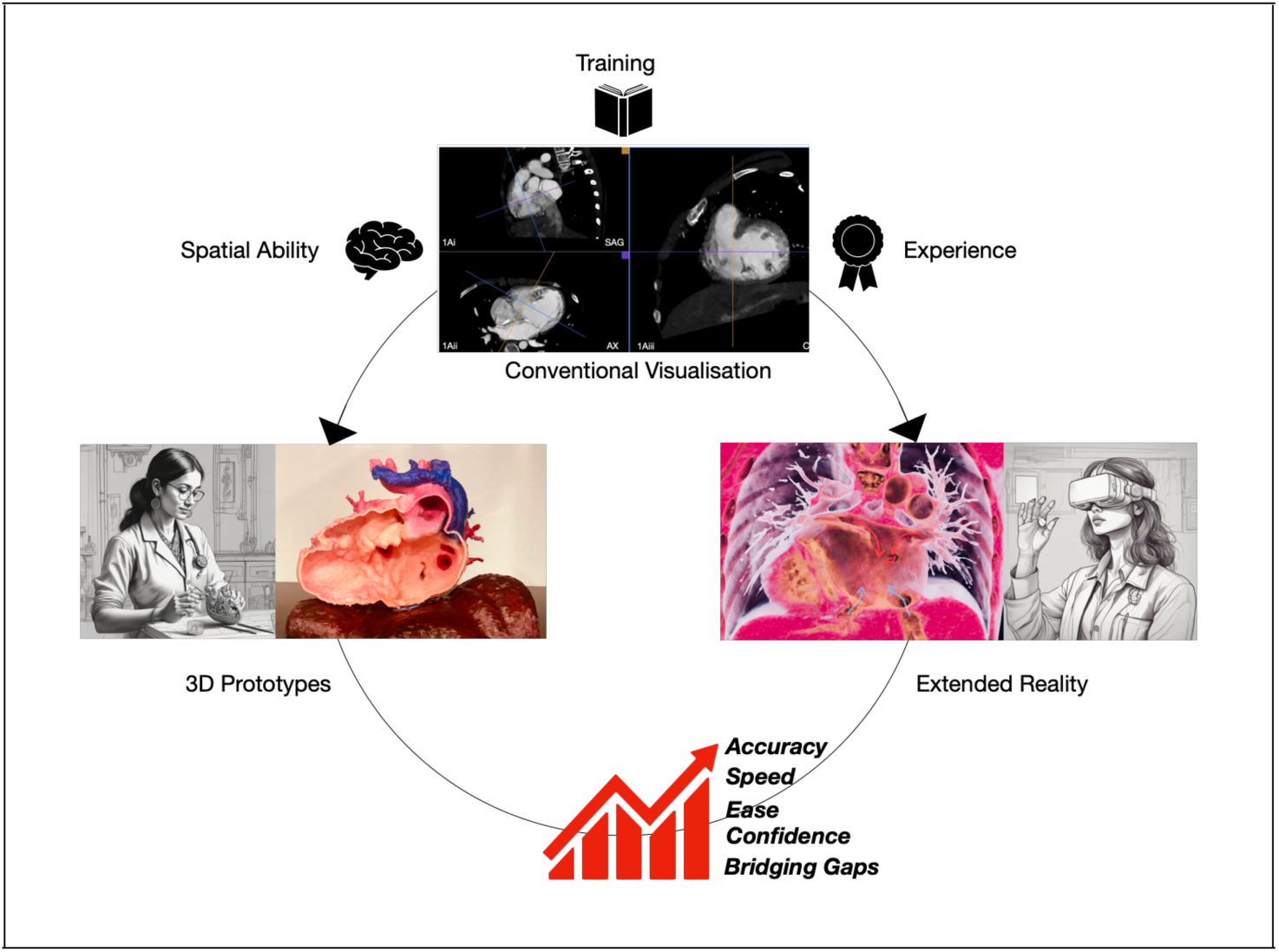

## Notes

### Competing Interest Statement

The authors have declared no competing interest.

### Funding Statement

This study did not receive any external funding.Partially funded by Institutional research grant of Amrita Institute of Medical Sciences

### Author Declarations

Ethics Committee of Amrita Institute of Medical Sciences, Kochi, India gave ethical approval for this work

### Summary of Updates

Word count reduced to 3000 words. Tables and figure legends edited.

