## Supplemental Table for "Immersive 3D Visualisation (3D Printing, Virtual Reality) enhances spatial understanding of complex congenital heart defects"

**Supplemental Table : Elements of segmental anatomy - Accuracy scores of Pediatric Cardiac consultants versus Trainees**

| SPATIAL UNDERSTANDING | | CDICOM | 3DP | VR | p-Value |
|---|---|---|---|---|---|
| **Ventricular relationship** | **Consultants** | 78.40% | 95.10% | 99% | <0.001 |
| | **Fellows** | 44.2% | 90.1% | 94.11% | <0.001 |
| | **Overall** | 68.6% | 94.11% | 97.3% | <0.001 |
| **Atrioventricular relationship** | **Consultants** | 77.40% | 96.40% | 93.90% | <0.001 |
| | **Fellows** | 58.03% | 93.05% | 85.7% | <0.001 |
| | **Overall** | 61.1% | 95.2% | 89.68% | <0.001 |
| **Ventriculo-Arterial relationship** | **Consultants** | 83.30% | 97.10% | 100% | <0.001 |
| | **Fellows** | 66.6% | 94.11% | 94.1% | <0.001 |
| | **Overall** | 77.1% | 96.04% | 98.03% | <0.001 |
| **Great Artery arrangement** | **Consultants** | 84.30% | 92.20% | 100% | <0.001 |
| | **Fellows** | 72.5% | 70.5% | 92.1% | 0.014 |
| | **Overall** | 81.04% | 84.9% | 96.6% | <0.001 |
| **Type/Location of Ventricular Septal Defect** | **Consultants** | 62.20% | 67.80% | 92% | <0.001 |
| | **Fellows** | 20% | 71.1% | 80% | <0.001 |
| | **Overall** | 48.14% | 68.8% | 87.4% | <0.001 |
| **Routability of Ventricular Septal Defect to Great Artery** | **Consultants** | 62.70% | 84% | 95% | <0.001 |
| | **Fellows** | 10.3% | 79.3% | 100% | <0.001 |
| | **Overall** | 45.5% | 81.3% | 96.6% | <0.001 |

| | | | | | |
|---|---|---|---|---|---|
| **Great Artery override** | **Consultants** | 36% | 58% | 42% | 0.302 |
| | **Fellows** | 8.3% | 33.31% | 33.31% | 0.264 |
| | **Overall** | 25% | 47.2% | 36.11% | 0.142 |

Table compares accuracy scores of pediatric cardiac consultants (cardiologists and surgeons) with pediatric cardiology fellows-in-training while interpreting specific elements of spatial anatomy. There was statistically significant improvement in accuracy of interpretation for both groups with use of immersive 3D formats, for all elements for spatial segmental anatomy. The lowest accuracy was in answering questions related to identification of overriding great artery - with statistically insignificant improvement even with 3DP and VR. This was possibly due to absence of pulmonary artery in some of the cases (either atretic or ligated during previous palliated surgeries). Difference between consultants and fellows was reduced with 3d formats.
